# A time-multiplexing approach to shared ventilation

**DOI:** 10.1101/2024.08.06.24311558

**Authors:** Jacob Q Yarinsky, Abby Blocker, Carolyna Yamamoto Alves Pinto, Christopher L Passaglia, Stefano Pasetto, Heiko Enderling, Aaron R Muncey

## Abstract

Ventilator shortages during the COVID-19 pandemic forced some hospitals to practice and many to consider shared ventilation, where a single ventilator is used to ventilate multiple patients simultaneously. However, the high risk of harm to co-ventilated patients secondary to the inability to treat anatomically different patients or safely adapt to dynamic ventilation requirements has prevented full adoption of multi-patient coventilation. Here, a time-multiplexing approach to shared ventilation is introduced to overcome these safety concerns. A proof-of-concept device consisting of electromechanically coupled ball valves to induce customized resistances and facilitate the delivery of alternating breaths from the ventilator to each patient is presented. The approach successfully ventilated two test lungs, and individualized tidal volume combinations of various magnitudes were produced. Over five hours of co-ventilation, consistency in tidal volume delivery was comparable to independent ventilation. Time-multiplexing was able to facilitate delivery of statistically unique tidal volumes to two test lungs and maintain the consistency of tidal volumes within each test lung while independently ventilated with identical parameters. The ability to adjust each test lung’s inspiratory pressures dynamically and independently was also demonstrated. The time-multiplexing approach has the potential to increase the viability of co-ventilation for ongoing and future ventilator shortages.

## Introduction

Surges of critical COVID-19 patients combined with a shortage of ventilators have forced healthcare providers to triage patients for survivability before ventilating to preserve resources [1], or attempt to ventilate multiple patients with a single ventilator [2,3]. Shared or co-ventilation has been used emergently in rudimentary forms without the capability to uniquely treat dissimilar patients, precisely adapt to continuously evolving ventilation requirements, nor monitor the ventilation of each patient [4–9]. While this practice can be temporarily lifesaving, current methods have the potential to cause serious harm from barotrauma or hyper- or hypoventilation [10,11]. This technique has been described by major medical societies, including the American Society of Anesthesiologists and Society for Critical Care Medicine, as unsafe and “should not be attempted because it cannot be done safely with current equipment [12].“

To improve the quality and efficacy of the technique, researchers have proposed and investigated possible solutions such as, anatomical patient matching approaches [2,3], 3D printing of patient-dependent flow resistors [7], and ventilator circuit manipulation [8–10]. These approaches have made significant strides in making co-ventilation a viable technique. Some of these innovations have even received Emergency Use Authorization from the Food and Drug Administration [13].

Initial attempts at co-ventilation have been described using simple ventilation splitting devices [4,5]. These devices demonstrated a ventilator’s ability to deliver tidal volumes large enough to satisfy the ventilation needs for at least two patients. However, the technique requires patients to have similar ventilation needs, lung size, and pulmonary compliance. The inability to allow patient-specific pressure adjustment requires clinicians to pair patients with similar anatomies and physiologies together, which is rarely feasible and assumes no changes in lung compliance or airway resistance throughout each patient’s disease progression [14,15]. Even minor variations in patient characteristics could lead to dangerous differences in delivered ventilation, leading to either low or elevated minute ventilation or barotrauma and an overall substandard and even dangerous level of patient care [15].

Different co-ventilation solutions account for this by adjusting the flow resistance induced in each branch of the split circuit [6–10]. These devices use Pressure Control Ventilation (PCV) and regulate the inspiratory pressures delivered to each patient by manually adjusting the effective radius of the individual patient circuits via clamps or valves embedded in the tubing. While an improvement over previously described splitters, the major drawback of these devices is their lack of precision while adjusting resistances, leading to the risk of inaccurate ventilation and patient harm. These devices also require additional monitors and sensors to assess the changes made and may not be intuitive for all hospital personnel. Furthermore, these devices do not address the risks of patient interaction by coughs, occlusions, and disconnections.

One co-ventilation circuit of interest proposed by Chase et al. is “in serial” co-ventilation [16], rather than “in parallel” approaches described above. While no physical prototype was developed by Chase et al., they conceived of an active solenoid valve that facilitates alternating breaths to each patient, dichotomizing the tidal volumes in the ventilator circuit at any given time. In response to this proposed co-ventilation technique, Freebairn and Park published a letter describing the limitations of the approach, including the approach’s inability to ventilate patients with a sufficiently high respiratory rate for patients with COVID-19 [17].

Here, we propose a time-multiplexing approach to co-ventilation, describe a proof-of-concept device, and demonstrate in test lung experiments that time-multiplexing can overcome the main obstacles of previously shared ventilation solutions, including prevention of air sharing with alternating breath delivery, quantitative pressure setting for each patient, inspiratory flow and tidal volume monitoring without additional sensors, and safe pressure adjustments in real-time. We describe time-multiplexing as an alternative approach to coventilation that can further increase the technique’s safety, expand access to life-saving ventilation in remote and underdeveloped areas, and provide a low-cost solution for future ventilator shortages.

## Materials and methods

Time-multiplexing is a signals communication technique that allows for transmission of multiple signals on a single data stream. The technique requires that the signals have finite bandwidth so they can be interdigitated in time without temporal overlap [18]. When applied to co-ventilation, this approach capitalizes on the biphasicity of the respiratory cycle which includes an inspiratory time (I-time) and an expiratory time (E-time). The I-time consists of the ventilator delivering the prescribed pressure or tidal volume. The E-time consists of a variable duration in which the patient exhales and the ventilator is at rest. With a timemultiplexing approach, the ventilator is set to produce breaths at twice the respiratory rate of a single patient, and rather than standby idly between two breaths of one patient, deliver a breath to another patient while one patient naturally exhales (Fig 1A). With a doubled PCV rate or Continuous Positive Airway Pressure (CPAP) mode, an external device can then take the ventilator inspiratory tube and, via a splitter, alternate, or multiplex, the airflow into the different tube branches for individual co-ventilated patients (Fig 1B).

**Fig 1.**
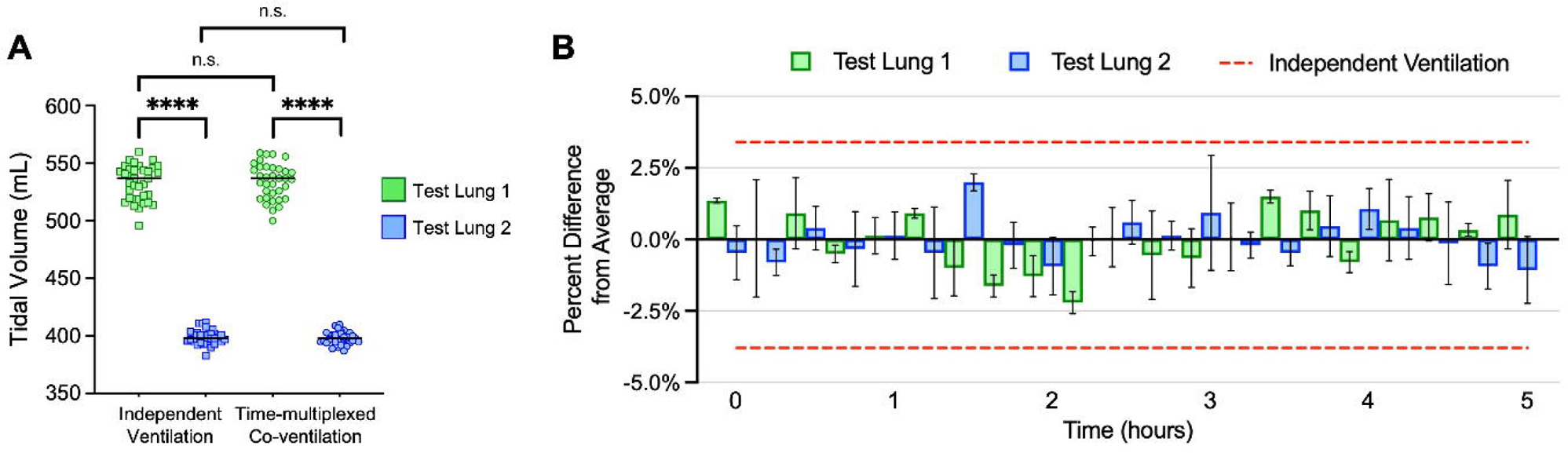
Schematic visualizations of the time multiplexing approach. (A) Schematic visualization of timemultiplexing with a 1:3 inhale-to-exhale ratio. The ventilation pause during the exhale phase of a single patient is used to ventilate an alternate patient with their own volume and pressure requirements. (B) Schematic visualization of a ventilator in either PCV or CPAP mode delivering air into the time-multiplexing device that alternates the airflow from a single ventilator to two co-ventilated patients with individual ventilation pressure prescriptions.

The use of Positive Pressure Ventilation (PPV) modes, such as PCV or CPAP, is key to this approach’s function. Unlike Volume Control Ventilation (VCV), PPV produces variable flow that is dependent on the resistance and compliance of the system it is ventilating. By ventilating patients at different times, the tidal volumes delivered to each patient are exclusively dependent on their individual airway resistance, lung compliance, and ventilation pressure. The goal of the proof-of-concept device presented here is to demonstrate the feasibility of the time-multiplexing approach by facilitating an alternating breathing pattern and inducing precise and dynamic flow resistances that allow delivery of unique tidal volumes.

The proof-of-concept prototype we have developed uses electromechanically coupled valves embedded in a ventilator splitting circuit, along with a user interface installed in a 3D printed housing (Fig 2). The circuit splitter was designed and 3D-printed to interface with standard medical equipment and minimize the effect of pressure build-up while allowing for enough space to fit the commercially available ball valves used in the device. Servo motors were connected to the ball valves using custom machined parts to fit the ball valves used in the circuit. Adapters were then attached to ball valves to interface with the continuation of the ventilator circuit. The electrical circuit of the device consists of a microcontroller, 2 servo motors, LCD screen, rotary encoder, 5 V regulator, and battery. The microcontroller is programmed to collect 4 parameters from the user including ventilator pressure, the desired pressures for each patient, and desired respiratory rate (I:E ratio) for the patients.

**Fig 2.**
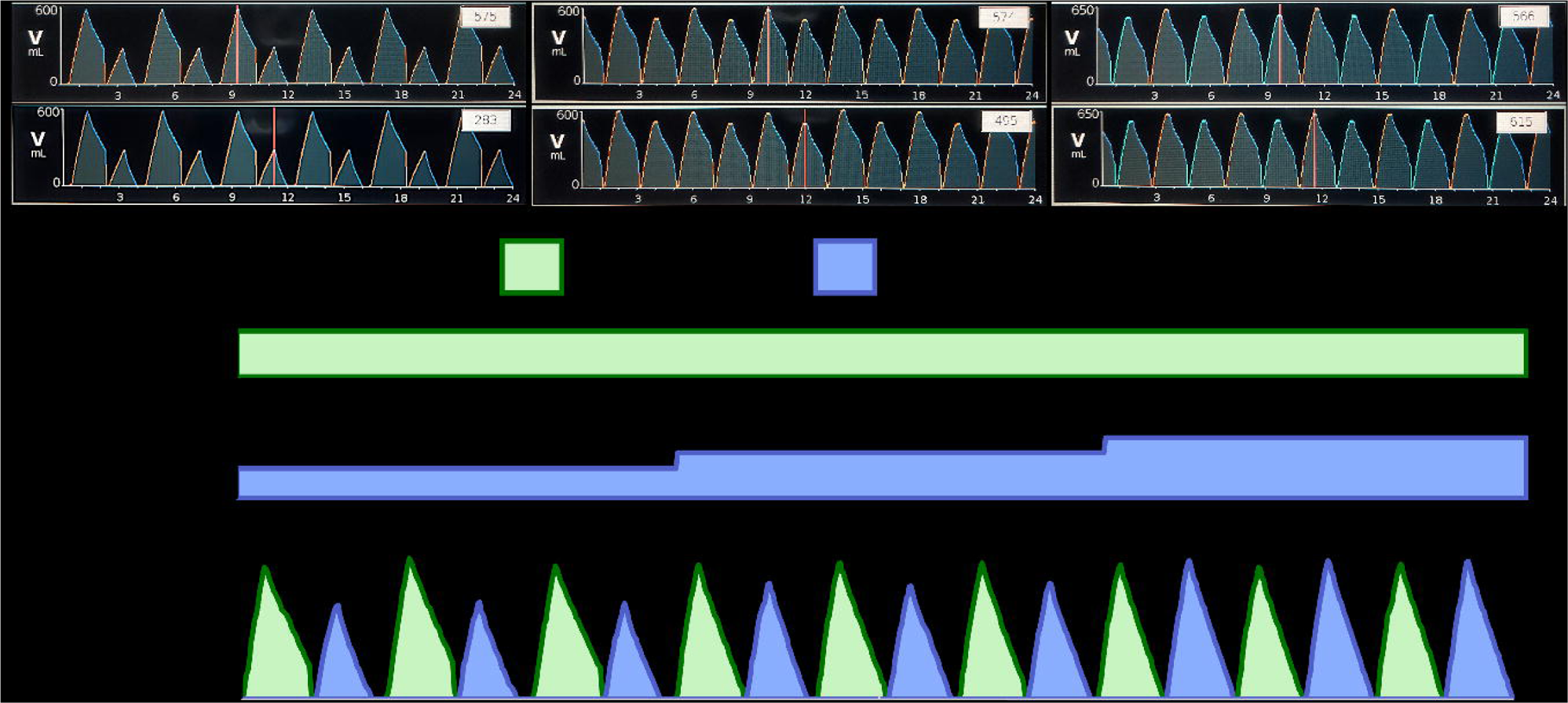
Various views of the time-multiplexing co-ventilation device prototype. (A) Isometric view of the prototype. Total dimensions are 6”L x 9”W x 4”H. User interface is shown on top of the device including rotary encoder knob, LCD screen, power switch, cooling fans and latches. (B) Inside bottom view with 3D printed splitter, PVC fittings, circuit adapters, ball valves, and servo motor connections. (C) Inside top view containing electrical components. Servo motors, batteries, microcontroller, and voltage regulator. All components were secured to the 3D printed housing with screws, and wires were soldered together.

Of importance, the motor-controlled ball valves in each branch of the device operate as both the solenoid valve and flow resistors. The use of servo motors enables the precise angular control of the ball valve to specific positions and can therefore be used to open and close each patient’s circuit, as well as partially open the valve to specific angles to induce precise resistances. The use of servo motor-controlled ball valves as flow resistors was employed for their ability to completely close and open over a short range of motion and time. Other valves, such as pressure limiting valves, can be more precise for pressure modulation, but actuating those valves to accomplish the temporal aspects of the time-multiplexing approach remains uncertain. Additionally, the rapid closing of the ball valve after each patient’s inspiratory phase prevents the sharing of contaminated gasses between patients, which has been shown to occur in other shared ventilation circuits leading to cross-infection risks and inconsistent tidal volume delivery [19].

The time-multiplexing approach requires the ventilator to work at double the respiratory rate of a single patient, and for the device to deliver alternating breaths to each patient. With the desired patient I:E ratio input to the device, a calculation is performed to determine the required ventilator respiratory rate. The expiratory time of the ventilator, *E_v_*, in seconds, is calculated by:

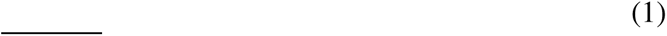

Where *I_P_* and *E_P_* are respectively the inspiratory time and the expiratory time of both patients in seconds. The microcontroller uses Equation 1 to accurately open and close the ball valves to maintain synchrony with the ventilator and to ensure that each patient’s inspiratory phase occurs within the other patient’s expiratory phase.

For example, to co-ventilate patients with a desired I:E ratio of 1:3 (15 breaths per minute, bpm), the ventilator must be set to deliver breaths at a 1:1 I:E ratio (30 bpm), using PCV. However, the maximum respiratory rate using PCV may be limited by the ventilator’s capabilities. For respiratory rates that exceed the ventilator’s maximum PCV rate, the CPAP mode of ventilation may be used. While the CPAP mode is not designed to deliver inspiratory pressures, the ventilator reacts to the opening of a valve and drives the pressure to the CPAP setpoint, imitating a pressure waveform similar to PCV. Using CPAP, the time multiplexing approach can facilitate a patient I:E ratio as low as 1:1 (30 bpm) by opening one patient’s valve while simultaneously closing the other. While used less often owing to risks of hyperventilation, auto-PEEP, among other anatomical and physiological impacts [20], the ability to produce high respiratory rates is presented here as a potentially useful feature of time-multiplexed co-ventilation. Common ventilator-patient I:E ratio combinations and the required ventilation modes are listed in Table 1.

**Table 1.** Required ventilation mode and I:E ratio for desired patient I:E ratio using time-multiplexing approach.

| Ventilator I:E Ratio Required for Desired Patient I:E Ratio |  |  |
| --- | --- | --- |
| Prescribed Patient I:E ratio (bpm) | Required Ventilator I:E ratio (bpm) | Ventilator Mode(s) |
| 1:5 (10) | 1:2 (20) | PCV or CPAP |
| 1:4 (12) | 1:1.5 (24) | PCV or CPAP |
| 1:3 (15) | 1:1 (30) | PCV or CPAP |
| 1:2 (20) | N/A* | CPAP Only |
| 1:1.5 (24) | N/A* | CPAP Only |
| 1:1 (30) | N/A* | CPAP Only |

\*Ventilators with the capability to use PCV at rates higher than 30 bpm can be used. Otherwise, the ventilator mode must be CPAP.

To quantitatively modulate the pressures delivered by the ventilator, the degree at which the motor-controlled ball valves open is correlated to a specific pressure drop using computational fluidic simulations in ANSYS Fluent (ANSYS 2021 R2, ANSYS STUDENT, Canonsburg, PA, USA). The CAD assembly of the ventilator circuit was imported and converted to a mesh using ANSYS SpaceClaim (ANSYS 2021 R2, ANSYS STUDENT, Canonsburg, PA, USA). The inspiratory pressure was set to 30 cmH_2_O at the inlet of the splitter, and the angle of the ball valve was adjusted in 10-degree increments between 0° and 90°. Turbulent flow was observed within the circuit, so the maximum (peak) pressure was sampled 1-inch after the ball valve (measurement plane) to eliminate the effects of eddies on the pressure measurements (Fig 3A). These data were used angle to correlate the flow resistances induced by the ball valves with the more clinically relevant effective inspiratory pressures. The simulation data (Fig 3B) were fit with the 3rd order polynomial

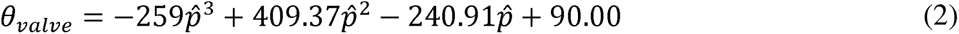

with high accuracy (R2 = 0.996), where *θ_valve_* is the angle of the valve during the inspiratory time of the patient in degrees, and *p̂* is the ratio between the effective inspiratory pressure for each patient to the pressure set on the ventilator. Additional visualizations of simulation data can be found in the supplement (Fig S1).

**Fig 3.**
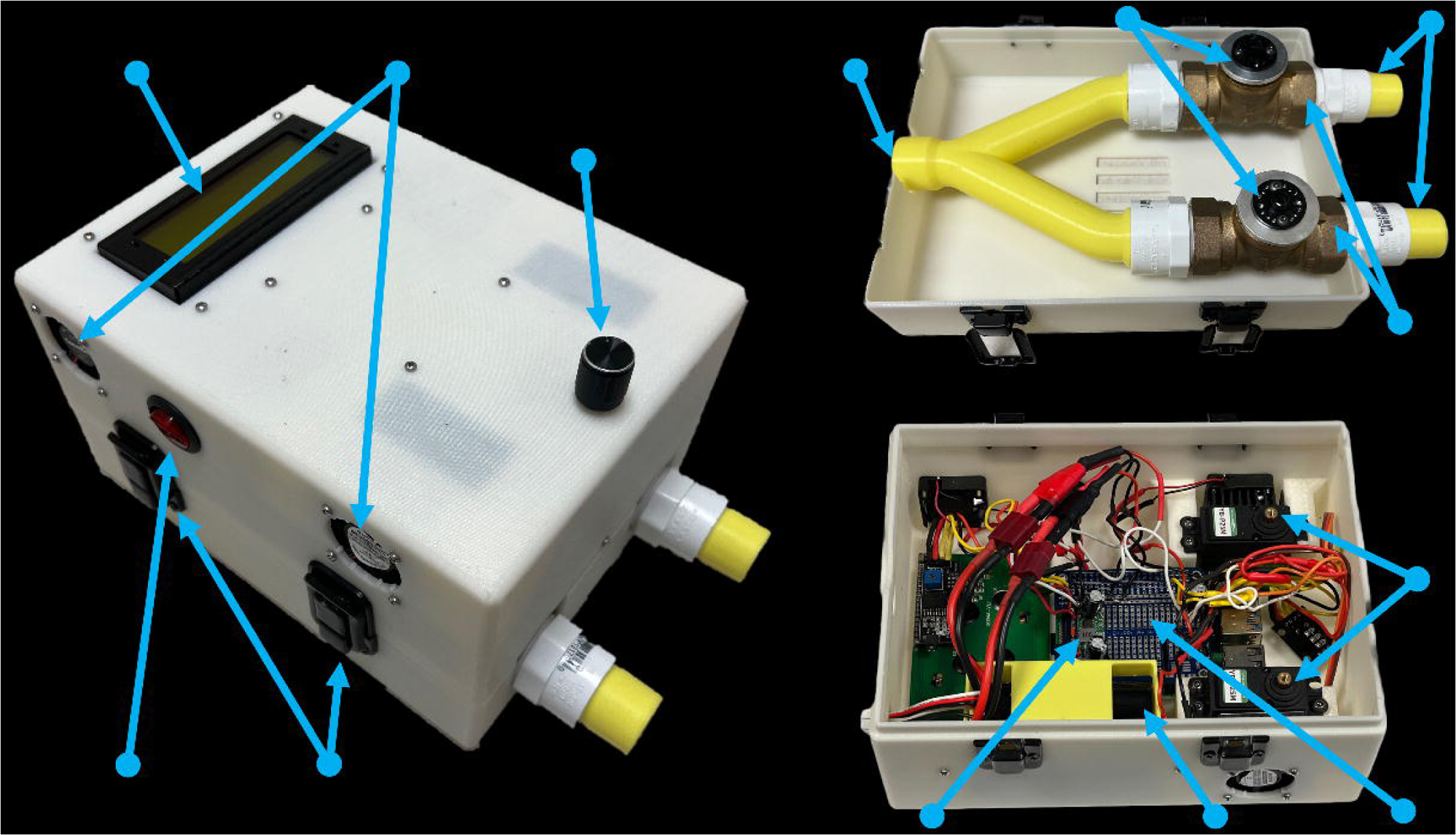
Calibration and validation of computational fluidic dynamic simulations to experimental data to correlate position of the servo motor to resistance induced by the coupled ball valves. (A) Mesh of CAD model of device ventilation circuit for 1 patient and front plane view of flow pathlines through circuit at ball valve angle of 20 degrees. (B) Simulation data fit with 3^rd^ order polynomial with R^2^=0.996. (C) Validated with experimental data with R^2^=0.986. panel. (D) Illustration of ventilation pressure waveforms delivered using the time-multiplexing device. The valves are closed in time zones A and C, allowing each patient to exhale regularly. In time zone B, the fully open ball valve during patient 1’s inspiratory period delivers the maximal pressure. In time zone D, the partially opened ball valve during patient 2’s inspiratory period delivers the prescribed (lower) pressure.

To experimentally validate the simulation results, the pressure drop caused by the partially opened ball valves was correlated to specific inspiratory pressures using the ventilators tidal volumes outputs. The tidal volumes delivered to a test lung with variable inspiratory pressures (set on the ventilator) and no resistance (ball valve completely open) were correlated with tidal volumes delivered at a single inspiratory pressure and variable ball valve resistances. The calibration curve (eqn. 2) was validated by the experimentally determined pressure (R2 = 0.986; Fig 3C). The experimental data were collected by directly referencing the inspiratory tidal volume output by the ventilator. The high agreement between simulated and experimental data supports the reliance on the ventilator’s inspiratory tidal volume as an accurate quantification of the ventilation provided by the device.

An illustrative example of integrating the alternating temporal pattern and ball valve calibration curve into the device prototype is shown in Fig 3D. The device opens and closes the ball valves to specific angles such that patient 1 receives 70% of the ventilator pressure (*θ_valve_*,*P*1 = 33°; eqn. 2) and patient 2 receives 100% (*θ_valve_*,*P*2 = 0°) in an alternating, time-multiplexed temporal pattern (eqn. 1) to co-ventilate both patients with an I:E ratio of 1:3.

To examine the functionalities of the time-multiplexing approach, test lungs of varying compliances, Venti Plus Adult Test Lung (low compliance; 0.01L/cmH_2_O) and MedLine Breathing Bag 2L (high compliance; non-linear), were co-ventilated with a single port Philips Respironics V60 BiPAP Ventilator. The ventilation circuit was constructed using clinical BiPAP ventilator tubing. The ventilator’s inspiratory tube was connected to the inlet of the device’s splitter. Each test lung’s circuit was connected to one of the two device outlets. Those circuits consist of an inspiratory and expiratory limb. The inspiratory limbs were connected directly to the different test lungs. The expiratory circuits were then connected to each other and allowed to exhale to room air. The expiratory circuits contained in-line PEEP valves. Bacterial/viral filters were placed in the circuit between the ventilator and the device as well as between the test lungs and the PEEP valves. For all experiments, ventilator PEEP was set to 4cmH_2_O (the minimum allowed by the ventilator). PEEP values reported below correspond to the in-line PEEP valves. Tidal volumes were collected directly from the ventilator screen and used to evaluate the ventilation delivered, as tidal volumes must be precisely controlled to maintain adequate oxygenation while preventing trauma [21].

We evaluated the time multiplexing co-ventilation device by assessing the accuracy and consistency of tidal volumes delivered to two test lungs to the tidal volumes of an independently ventilated test lung. We further evaluated various co-ventilation tidal volume combinations that cover a wide range of ventilation needs, and the ability to independently adjust the ventilation to a single lung without affecting ventilation to the other. Statistical analysis was performed to analyze the differences between the tidal volumes delivered to both test lungs via independent ventilation and time-multiplexed co-ventilation. An unpaired t-test with Welch’s correction for uneven standard deviations was performed for each comparison. Independent ventilation was performed with Pressure Control Ventilation provided by the same Philips Respironics V60 ventilator through the same ventilation circuit (with no flow resistance induced by the ball valve).

## Results

The ability to set specific inspiratory pressures for each patient with time-multiplexing was tested by comparing the tidal volumes delivered to a Medline Breathing Bag 2L (test lung 1) and a Venti Plus Adult Test Lung (test lung 2) when being ventilated individually and co-ventilated with identical parameters. Test lungs were ventilated with a ventilator inspiratory pressure set to 20 cmH_2_O, respiratory rate of 20 bpm (patient rate = 10 bpm), and resistances that corresponded to effective inspiratory pressures of 12 cmH_2_O and 16 cmH_2_O for test lungs 1 and 2, respectively. In-line Positive End Expiratory Pressure (PEEP) valves were set to 5 cmH_2_O for both lungs. Inspiratory tidal volumes were recorded for each test lung (N=35 breaths) directly from the ventilator screen. Test lungs 1 and 2 received an average tidal volume of 533mL (+/− 15.2 mL standard deviation) and 399mL (+/− 6.2 mL), respectively (Fig 4A). Time-multiplexed co-ventilation delivered comparable tidal volumes to both test lungs (534 +/− 15.3 mL and 398 +/− 5.4 mL). Both independently ventilated and and co-ventilated test lungs received significantly different tidal volumes (p < 0.0001) with different pressure settings, but, the tidal volumes delivered to test lung 1 and test lung 2 in independent and co-ventilated settings were not statistically significant (p = 0.72 and p = 0.39, respectively). These results demonstrated that, similar to independent ventilation, the time-multiplexing device prototype can induce test lung-specific inspiratory pressures and that the calibrated resistances enable accurate inspiratory pressure setting.

**Figure 4.**
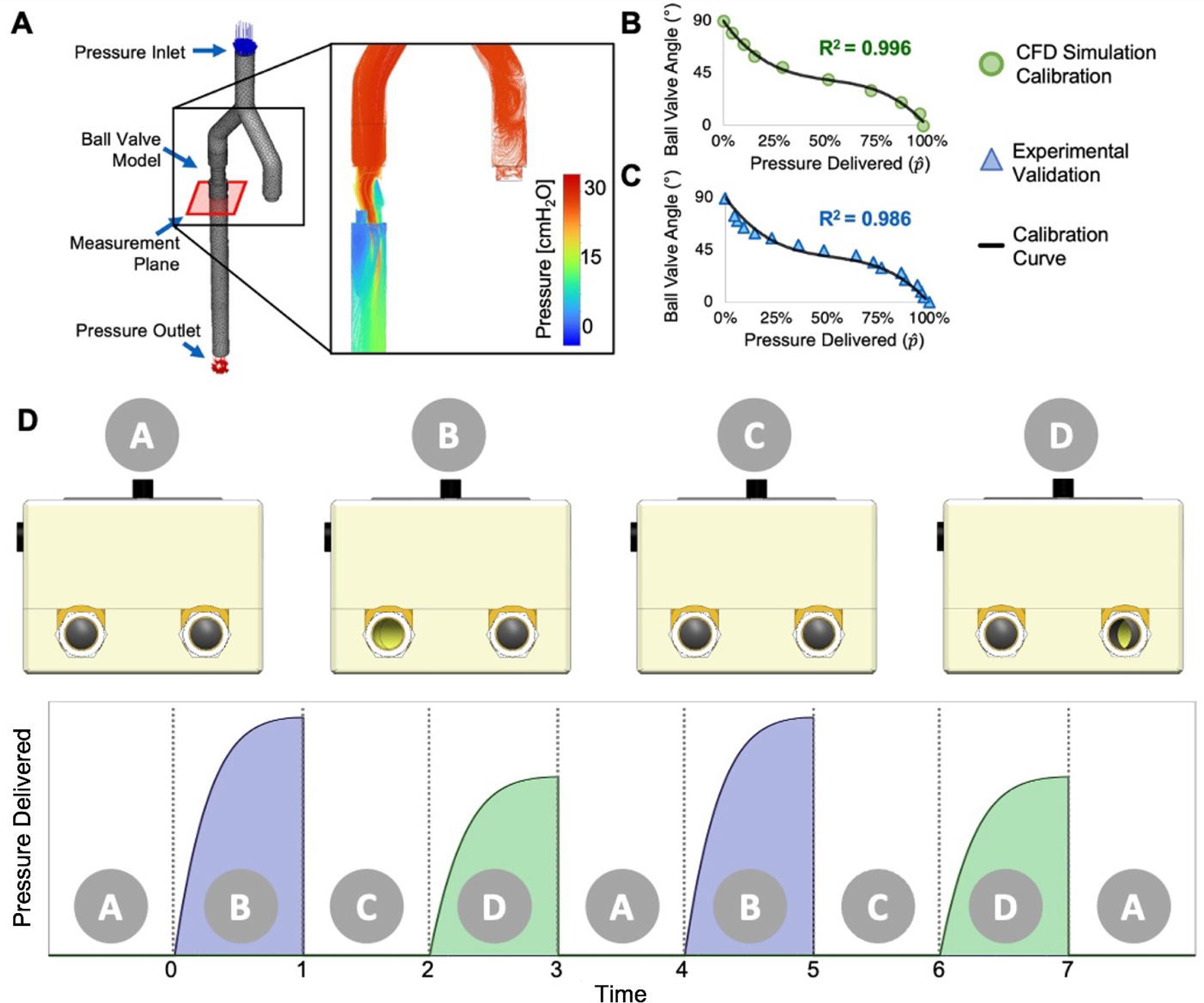
Comparison of time-multiplexed co-ventilation to independent ventilation. (A) Timemultiplexed co-ventilation of two test lungs with different tidal volumes provides similar tidal volumes compared to individual ventilation of each test lung. (B) Variability of delivered tidal volumes to test lungs over 5 hours of continuous co-ventilation did not exceed the variability of tidal volumes delivered to an independently ventilated test lung (red dashed lines). The mean of 3 tidal volume measurements at each time point with standard deviation error bars is shown for each time-multiplexed co-ventilated test lung.

The consistency of time-multiplexed co-ventilation was assessed by analyzing tidal volumes delivered over 5 hours of continuous individual or co-ventilation. Three tidal volumes were sampled every 15 minutes and collected directly from the ventilator screen (n=63 per test lung). For the co-ventilation, the inspiratory pressure was set to 30 cmH_2_O on the ventilator, and then set to 20 cmH_2_O and 30 cmH_2_O on the device for test lung 1 and test lung 2, respectively. In-line PEEP valves were set to 5 cmH_2_O for both lungs. Tidal volumes delivered to the test lungs were consistent throughout the experiment. The average tidal volume for test lung 1 was 500 +/− 5.5 mL, a variation of 1.1%. For test lung 2, the average tidal volume was 682 +/− 8.6 mL (1.3%). Individual test lung ventilation resulted in maximum tidal volume variation between +3.4 and 3.8% of the average tidal volume (Fig 4B). All time-multiplexed breaths delivered by the ventilator were within 3% of each test lungs average tidal volume. Therefore, time-multiplexed co-ventilation did not increase the variation of tidal volumes delivered to each patient compared to independent ventilation.

In contrast to co-ventilation approaches in which patients share breaths, a major clinical benefit of timemultiplexing is the ability to collect actionable patient-specific inspiratory tidal volume data directly from the ventilator screen. Since each patient is receiving an individual mechanical breath from the ventilator, the tidal volumes delivered for each patient are distinct and identifiable. This enables reliable, real-time, monitoring of the patient’s time-multiplexed ventilation. Using a ventilator with a respiratory rate of 30 bpm, inspiratory pressure of 30 cmH_2_O, and PEEP of 5 cmH_2_O, pressure settings to two test lungs were adjusted on the device to achieve different tidal volume combinations (Fig 5A). The full ventilator output, including pressure, flow, and volume waveforms, is shown in Fig S2.

**Figure 5.**
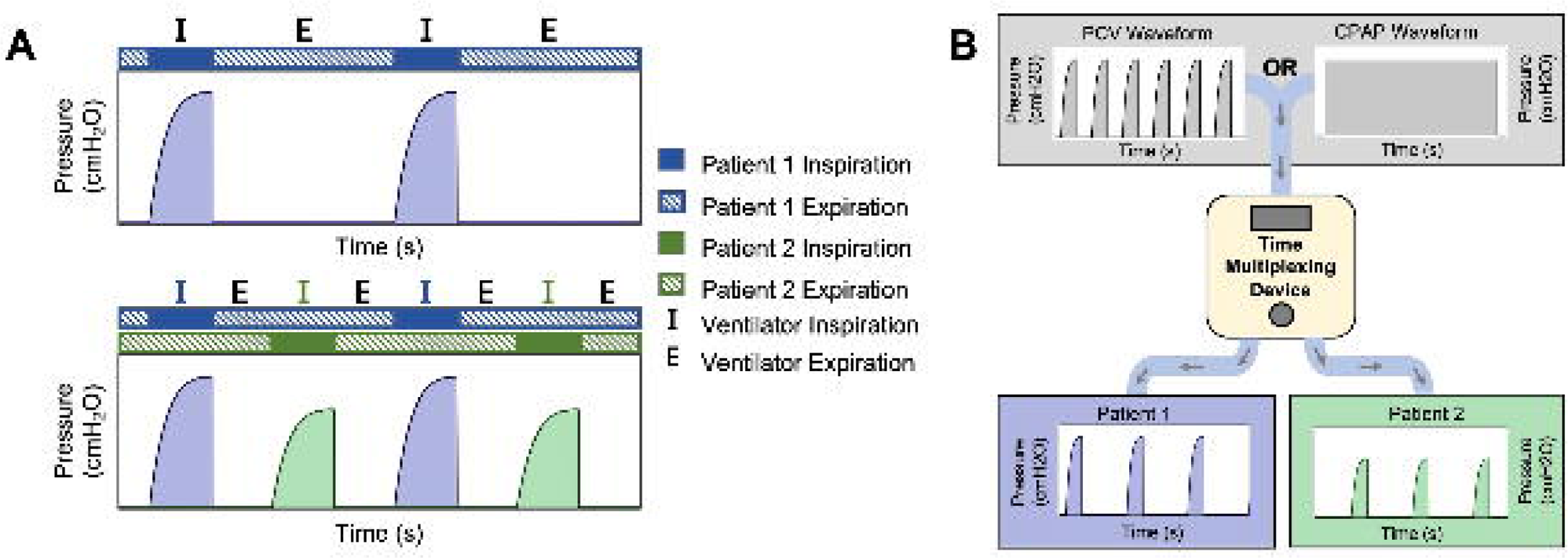
Various tidal volume combinations obtained and monitoring of independent adjustments. (A) Tidal volume readouts displayed on the ventilator during a single co-ventilation session with ball valve positions adjusted dynamically to achieve various tidal volume combinations. Left to right: Tidal volumes of approximately 575mL and 283mL breaths to each test lung. Tidal volumes of approximately 574mL and 495mL breaths to each test lung. Tidal volumes of approximately 566mL and 615mL breaths to each test lung. (B) Inspiratory pressures to one test lung were independently changed during co-ventilation and no impact on tidal volumes for the other test lung were observed.

Finally, time-multiplexing allows users to dynamically adjust the pressure delivered to one patient without impacting the ventilation of the other. The ventilator was set to PCV mode at a pressure of 25 cmH_2_O and a respiratory rate of 20 bpm, and pressure settings to test lungs 1 and 2 were set to 15 cmH_2_O and 10 cmH_2_O, respectively. The ventilator screen was paused every 18 seconds to collect tidal volume data, at which time the pressure setting for test lung 2 was increased by 5 cmH_2_O, and then the ventilator screen resumed data output. This process was repeated twice until the pressure setting of test lung 2 was equal to 20 cmH_2_O (Fig 5B). During the evaluated 54 seconds, consistent tidal volume delivery to test lung 1 is seen despite dynamic quantitative pressure changes to test lung 2.

## Discussion

Herein, we described a time-multiplexing approach to co-ventilation that could overcome some of the limitations of previously described co-ventilation solutions. While time-multiplexing has a long history in other applications, this is the first experimental application of the approach to assist mechanical ventilation of multiple simulated patients connected to a single ventilator. In the limited combinations of lung compliance, resistances, and ventilation settings tested, we demonstrated that time-multiplexing could address the monitoring and precision drawbacks of simple splitter and resistive splitter devices developed early in the COVID-19 pandemic. We demonstrated the ability to set individual quantitative inspiratory pressures for both patients, similar to how a clinician would set the inspiratory pressure for a patient being ventilated independently. Combined with adjustable in-line PEEP valves, the driving pressure for each patient was controlled. The temporal aspect of time-multiplexing unlocks the ability for clinicians to monitor the tidal volumes delivered to each patient without additional flow monitors or sensors and allows dynamic adaptation of ventilation parameters for an individual patient without impacting the other co-ventilated patient. These capabilities are crucial for clinicians to make informed decisions for critically ill patients who require individualized care.

While co-ventilation is generally reserved for emergencies, time-multiplexing with an auxiliary device provides an opportunity for ventilators to deliver personalized ventilation to multiple patients in nonemergent scenarios, too, such as low-resource settings, developing nations, or military applications.

Time-multiplexing has clear advantages, yet, in its current form, the approach is limited as it requires an accessory device that modulates a ventilator’s airflow rather than generating the airflow itself. One major limitation of the current implementation is the loss of expiratory flow data. The overlapping ventilation waveforms of each patient obscure the detection of auto-PEEP, prolonged exhalation, or spontaneous breathing efforts. The waveform output of the ventilator is driven by internal algorithms, which allow continuity of waveforms from breath to breath, but expiratory data readouts from the ventilator may be inaccurate with the external device attached.

Another limitation is the lack of an inspiratory trigger. Removing the patient’s ability to trigger a breath is required to maintain synchronicity with the ventilator’s respiratory rate. This also further limits the applicable ventilation modes that can function with the approach. Ventilation modes that require spontaneous breaths, such as Pressure Support Ventilation, cannot be used. The approach also requires both patients to be treated with the same ventilation mode and have identical I:E ratios and FiO_2_ concentrations, but there are avenues to address these limitations as discussed below.

The requirement for identical I:E ratios is based on the capabilities of the ventilator. A ventilator is programmed to deliver breaths at a uniform rate, which limits the approach’s ability to ventilate patients with different I:E ratios using the PCV mode. However, using the CPAP mode, a small subset of unequal I:E ratio combinations can be achieved using this approach. These combinations are discussed further in the supplement (Fig S3). Furthermore, the temporal pattern could be programmatically manipulated to allow for mismatched I:E ratios depending on the desired I:E ratios of the patients. With the advancements made in parallel co-ventilation, one may use this approach to co-ventilate with any I:E ratio combination, including inverse I:E ratios [17], while parallel co-ventilation temporarily occurs as the breathing patterns phase in and out of synchrony. Time-multiplexing can theoretically be used to co-ventilate more than two patients by integrating more branches and motor-controlled valves. However, this would also limit the maximum respiratory rate and I:E ratios available for each patient.

There are also many opportunities to improve the time-multiplexing approach to co-ventilation. The temporal isolation of each patients’ breaths presents a possibility to use VCV as the imbalance of pulmonary characteristics would no longer impact flow distribution. PCV has been favored over VCV specifically in coventilation for its decreased risk of ventilator-induced lung injury, yet VCV offers specific control of tidal volumes that may be beneficial for co-ventilation of patients with similar tidal volume needs and extremely mismatched pulmonary characteristics. Use of time-multiplexed co-ventilation with VCV remains dependent on a ventilator mode’s ability to produce time-cycled tidal volumes of alternating magnitude. Similarly, patient specific FiO_2_ could be achieved by relying on a ventilator to dynamically control the gas mixing in an alternating pattern.

Considerations for cost are also important with the development of low-cost ventilators [22,23]. The use of electrical components increases cost compared to plug-and-play mechanical components and would prolong the route to regulatory approval for clinical applications. While the time-multiplexing approach requires active components, the current design competes financially with low-cost ventilators and ventilator stockpiling. As seen during the COVID-19 pandemic, supplying medical and electrical components during times with ventilator shortages is a complex issue. Therefore, changes to a time-multiplexing device are probable to increase the approach’s viability for clinical translation.

The approach and prototype as currently described can directly interface with inexpensive CPAP machines. Any CPAP machine that can produce sufficiently high pressures and output tidal volume data can be adapted into a co-ventilation machine using the device. It is also possible to incorporate time-multiplexing as a unique mode into conventional ventilators with the associated software, ports, and mechanical components. Such integration could increase its compatibility with sensors and alarms, oxygen fraction control, different ventilation modes, and eliminate the limitations of an external device’s battery life, and new user interface.

While herein we demonstrated proof-of-concept, time-multiplexed co-ventilation must be further tested to comply with FDA regulations, given its novel approach to ventilation. Obtaining FDA/MDR approval will require more rigorous studies to ensure targeted tidal volumes and inspiratory pressures are delivered and a thorough evaluation of the device’s efficacy and safety in use cases that represent the myriad of possible clinical presentations. Future studies may include animal experiments using a porcine lung model and experiments using advanced lung simulators. Independently validating tidal volume, pressure, and flow measurements and dynamically adjusting respiratory mechanics, such as pulmonary compliance and airway resistance will be necessary to properly model the ventilation of a patient. Testing with different ventilators is also required to analyze the approach’s generalizability. Emulating the studies performed with other coventilation devices would be extremely valuable in analyzing this approach’s clinical benefit [9,24].

Questions remain regarding the technique of co-ventilation. There is a lack of clarity of when, if, or how the technique should be employed. The COVID-19 pandemic has brought about a plethora of innovation in the field of shared mechanical ventilation that has the ability to change the course of future ventilation shortages. The time-multiplexing approach to co-ventilation described here is a novel contribution to the field and may offer a viable, safe, and cost-effective option for multi-patient co-ventilation.

## Data Availability

All raw data files are available from the google drive database: https://drive.google.com/drive/folders/1eeTADDik8wftpbh137GedWNj5aAFqwQh?usp=sharing

## Acknowledgements

We thank Kim DeRose, Bridget McCormick, Kelly Belger, and the staff members of the Anesthesiology and Respiratory Therapy Departments at Moffitt Cancer Center who provided clinical insight, ventilators, test lungs, and workspace throughout the project, while also caring for patients amidst the COVID-19 pandemic. We also acknowledge Leila Sorrells, Cristian Benito Elias, Jazmine Enderling, and the Design for X Laboratory of the University of South Florida for their respective support and assistance in 3D printing, computational fluidic dynamic simulations, data collection, and prototyping.

## Funding and conflicts of interests

This work was supported by the H. Lee Moffitt Cancer Center and Research Institute Department of Anesthesiology Discretionary Fund and a BioEngineering at Moffitt pilot award. All authors are listed as inventors on an international patent application for a time-multiplexed co-ventilation device prototype (PCT/US2022/025218). This does not alter our adherence to policies on sharing data and materials.

