## supplement for "A time-multiplexing approach to shared ventilation"

**S1 Fig. Additional pressure pathline profiles of CFD simulations.** 0-, 20-, 50-, and 70-degree positions shown. Peak pressure recorded in ANSYS software for use in CFD calibration curve (Fig 3B).

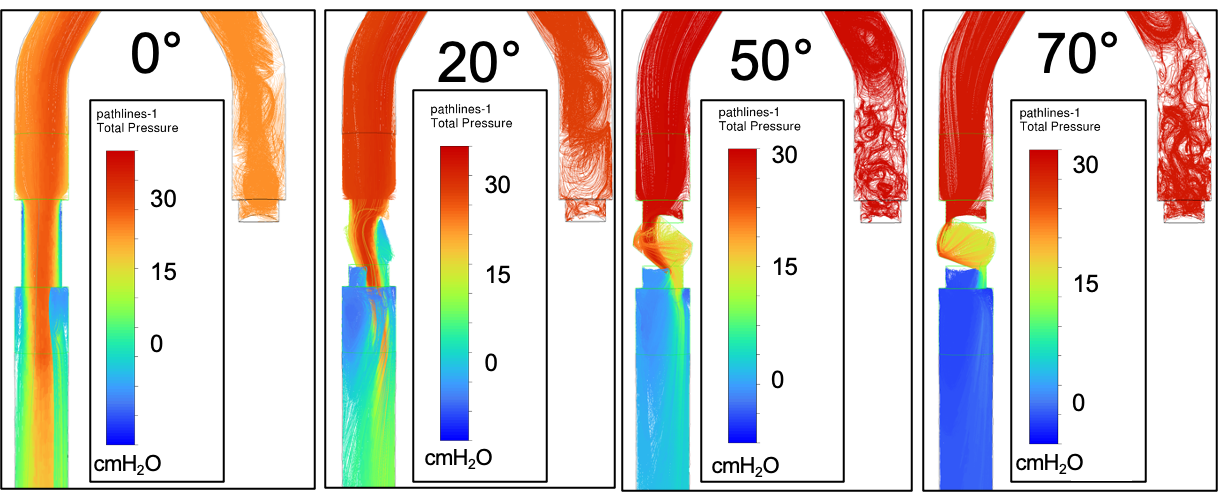

**S2 Fig. Full ventilator screen readout at various volume combinations.** Full pressure, flow, and tidal volume waveforms output by the ventilator during time-multiplexed co-ventilation as seen in Fig 5A. Ventilator set to respiratory rate of 30 bpm (I:E = 1:1; Patient RR = 15bpm, I:E 1:3). Expiratory flow and volume waveforms are driven by internal algorithms and reliance on their data is not possible with the device as constructed. Volume combinations of 575-283, 574-495, 566-615 shown with test lung 1 and 2 respectively.

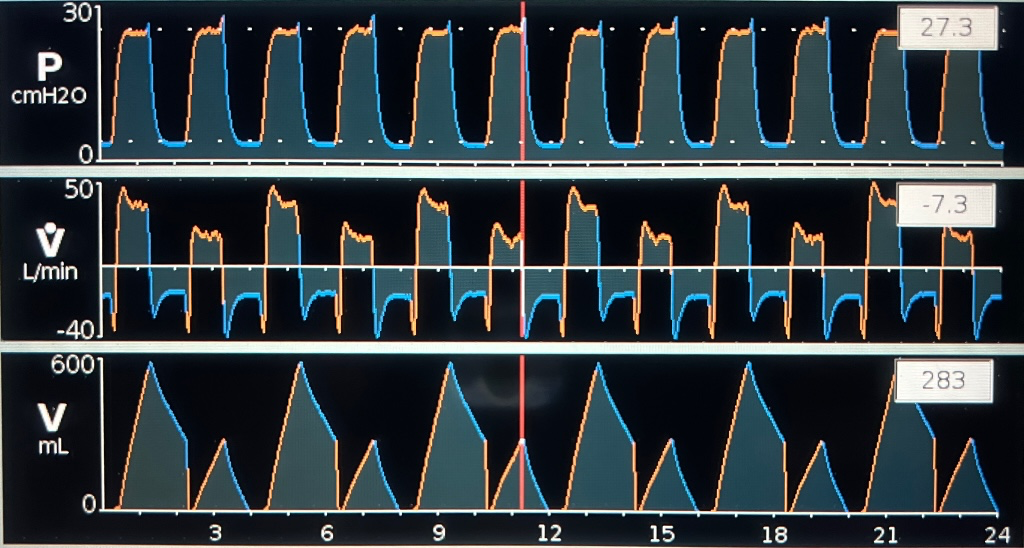

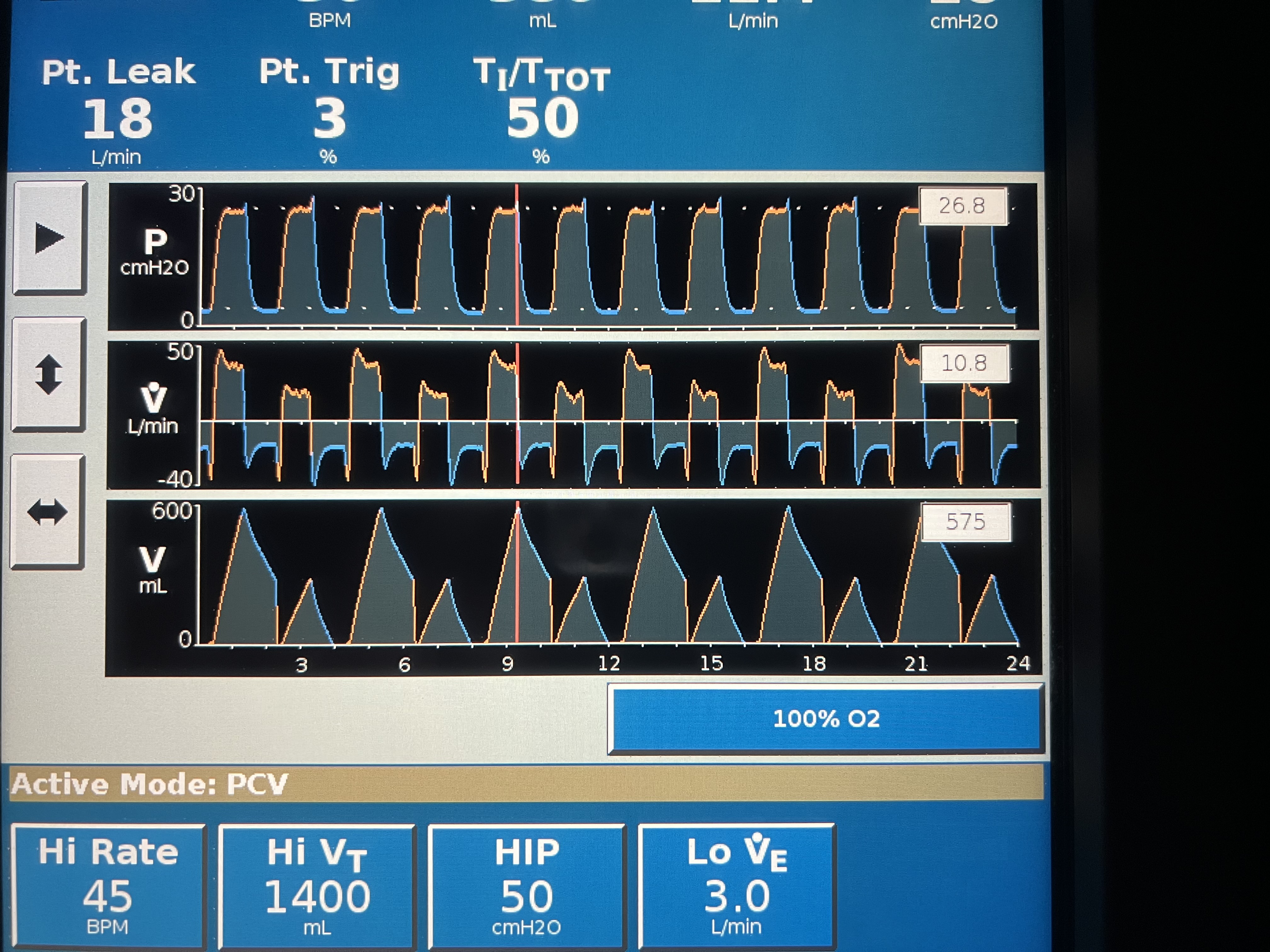

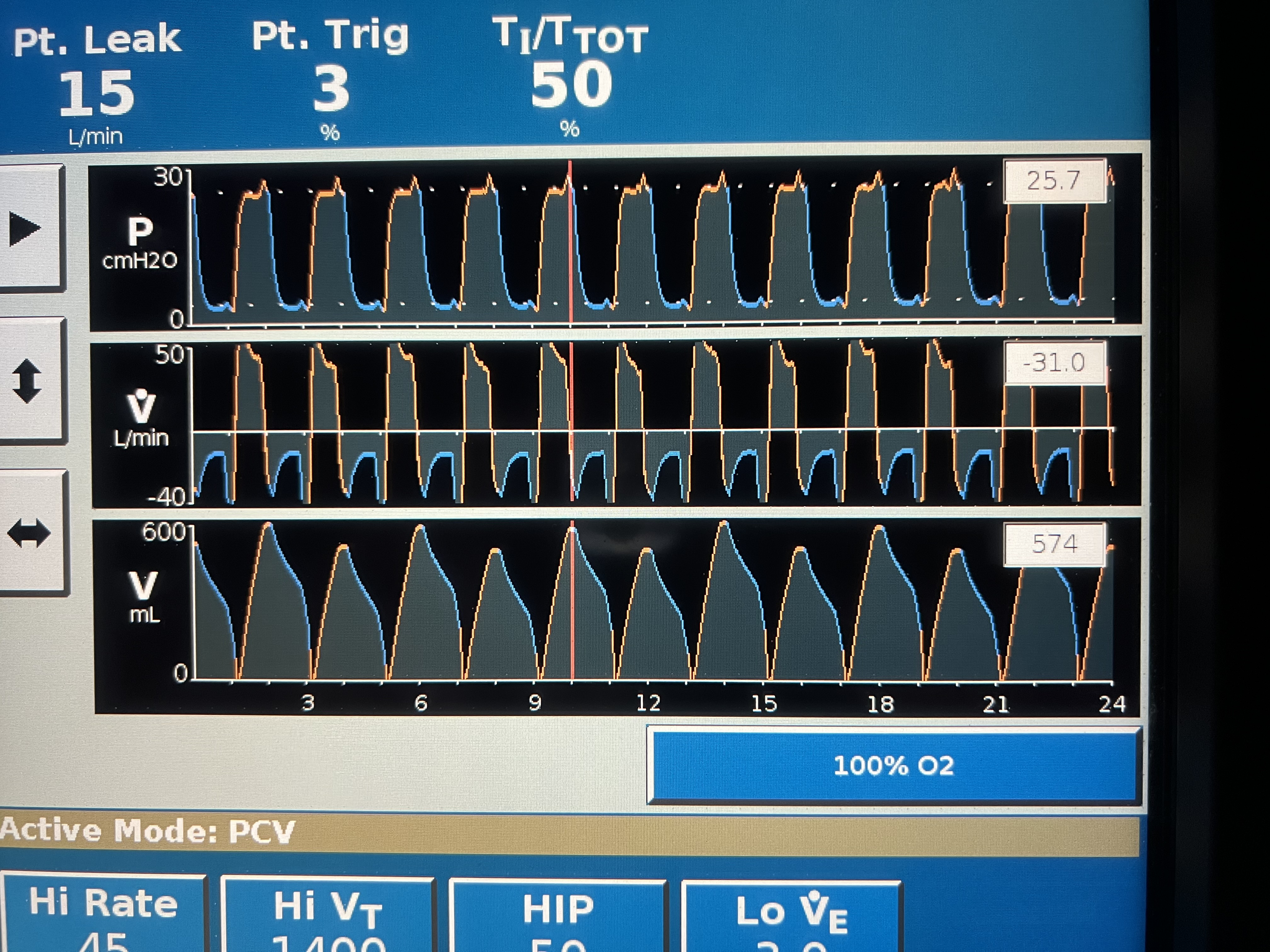

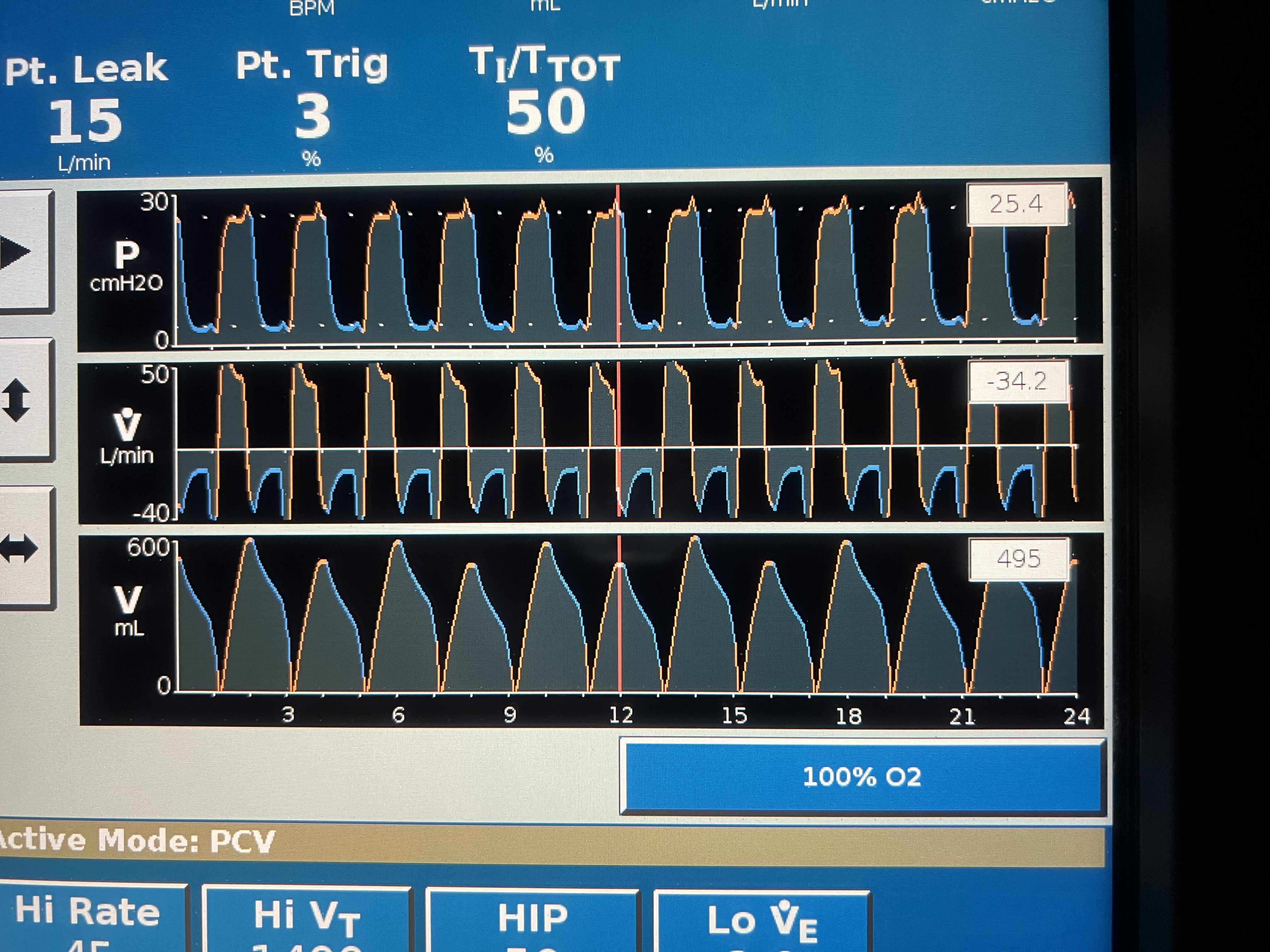

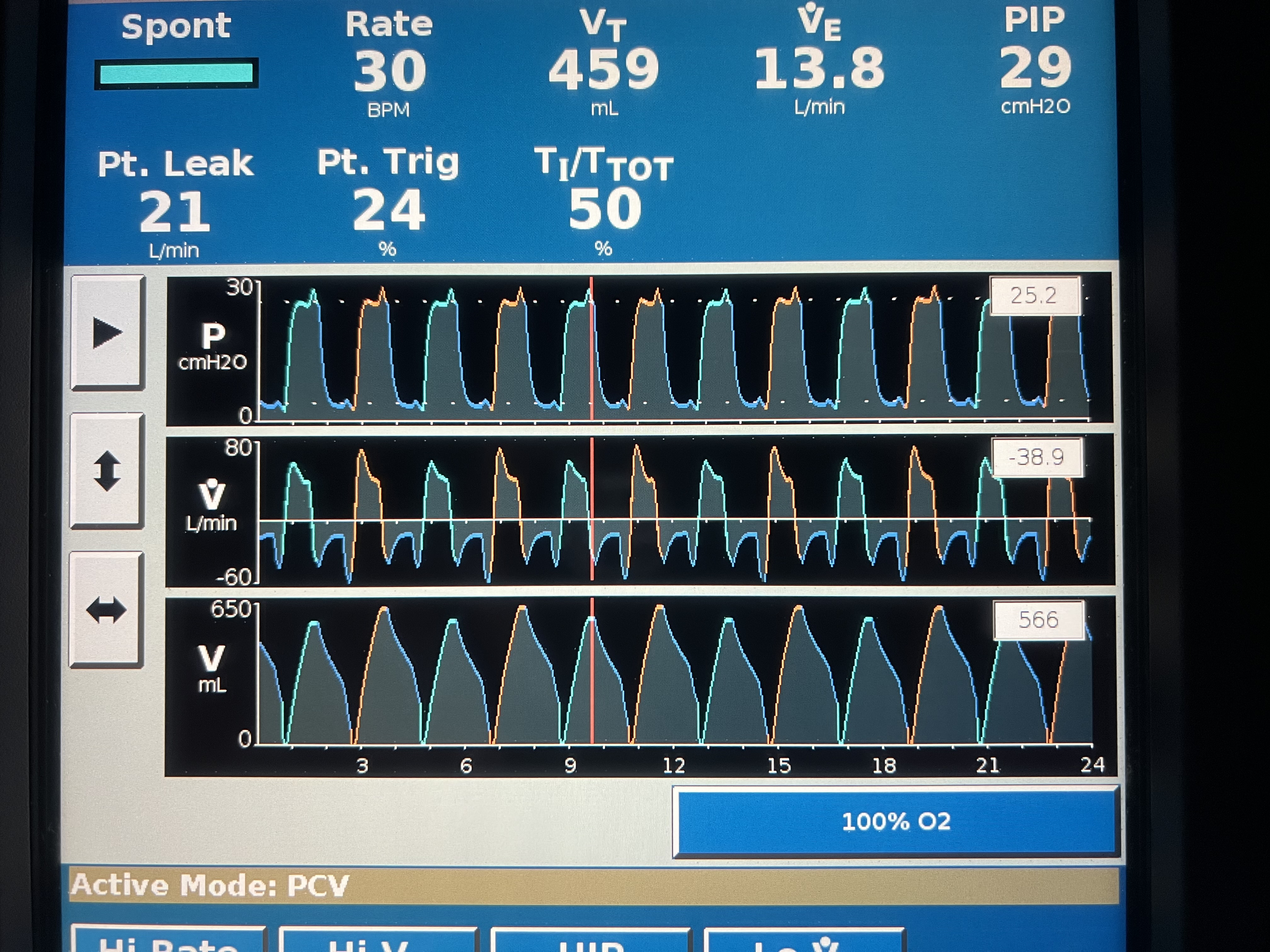

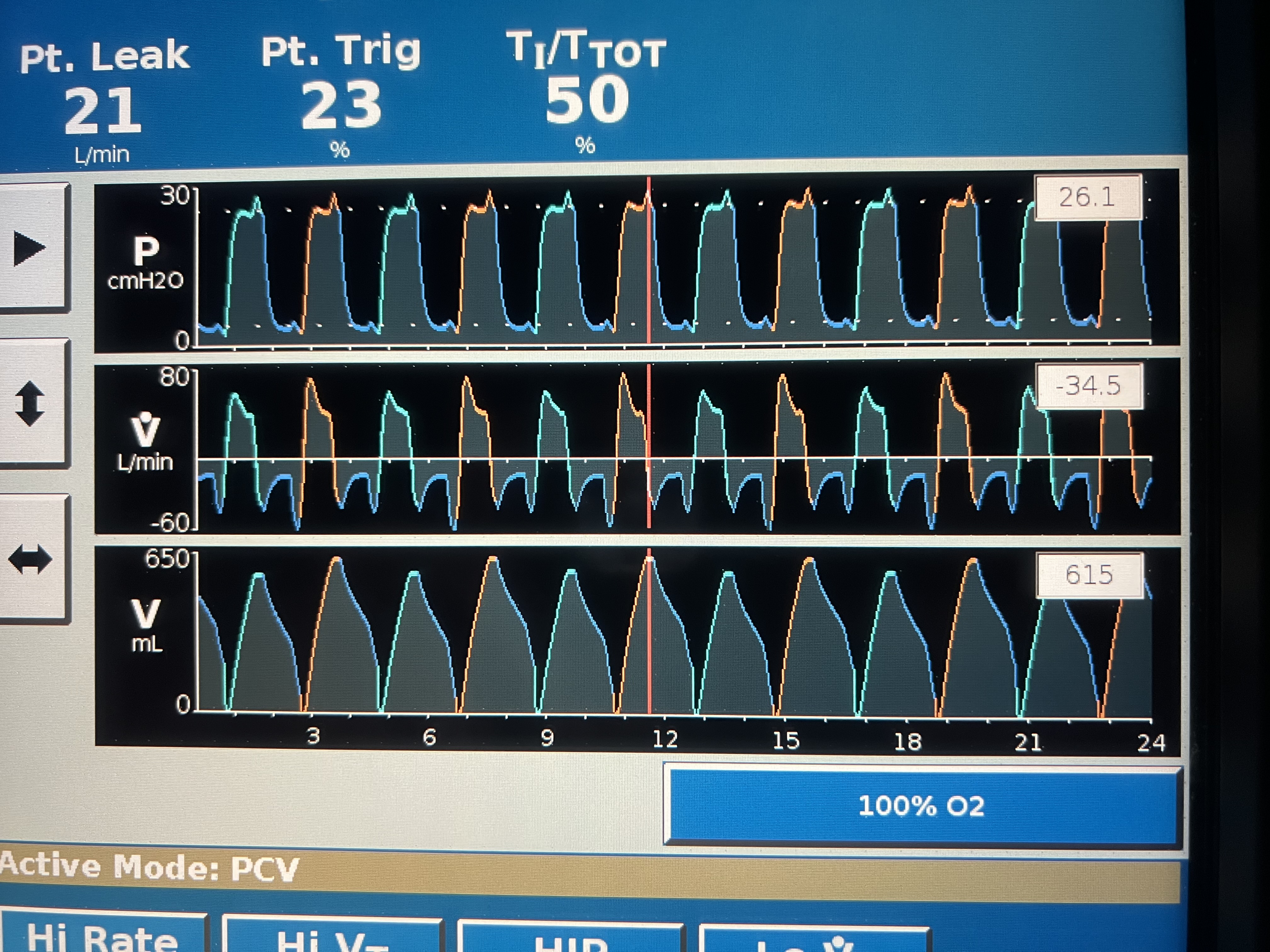

**S3 Fig.** **Mismatched I:E ratio combinations that do not temporally overlap with time-multiplexed co-ventilation.** Potential combinations were limited to a respiratory rate >10 breaths per minute and E-time multiples of 0.5 seconds. I:E combinations take the form: $T_{E,P_{2}}=2N\left( T_{E,P_{1}} \right)+1$, where $T_{E,P_{1}}$ is the expiratory time of patient 1, $T_{E,P_{2}}$is the expiratory time of patient 2, N can be any of the positive integers, and $T_{E,P_{2}}>T_{E,P_{1}}$. Schematic visualizations of I:E pairs of 1:1 and 1:3, 1:1 and 1:5, 1:1.5 and 1:4, and 1:3 and 1:5 are shown.

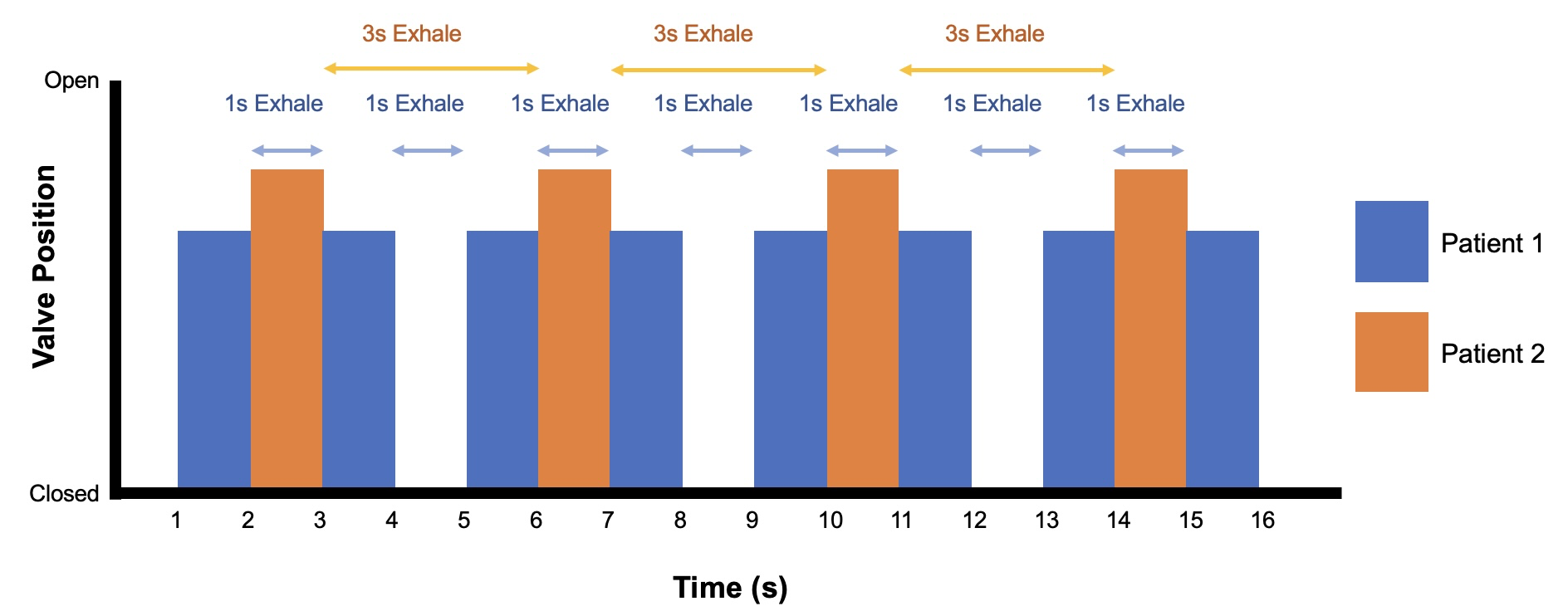

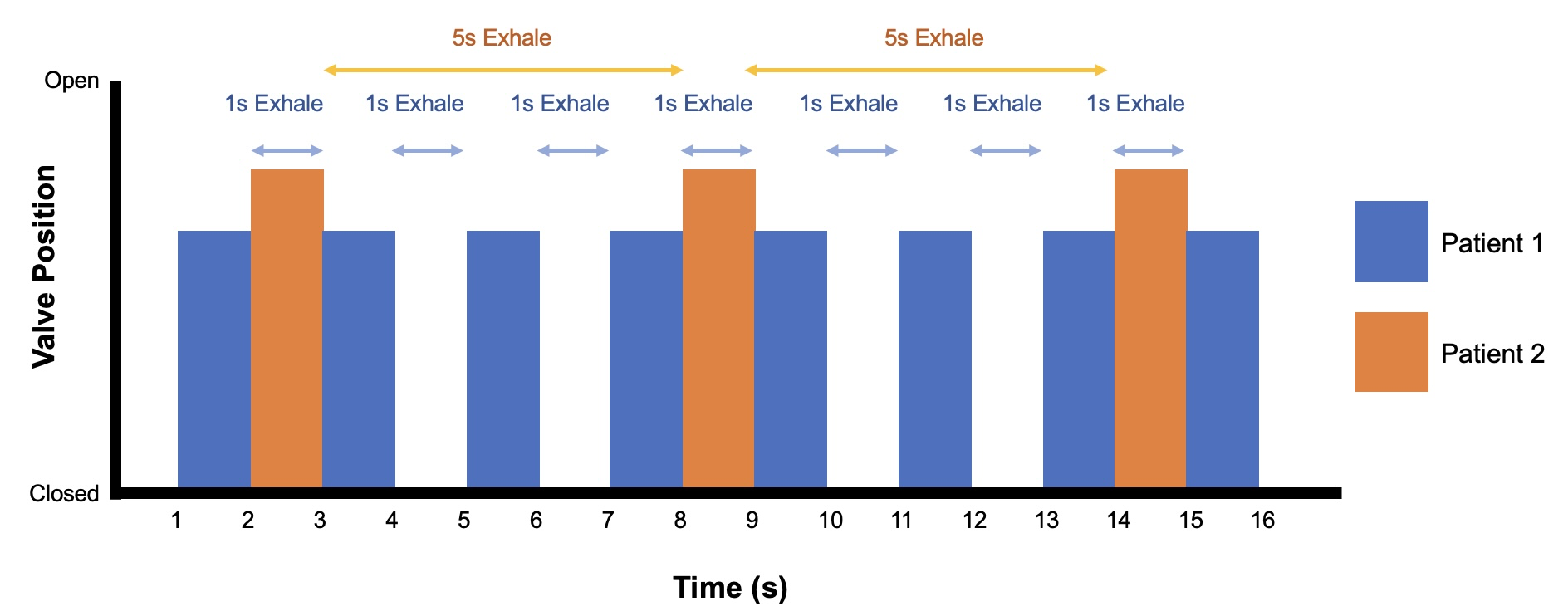

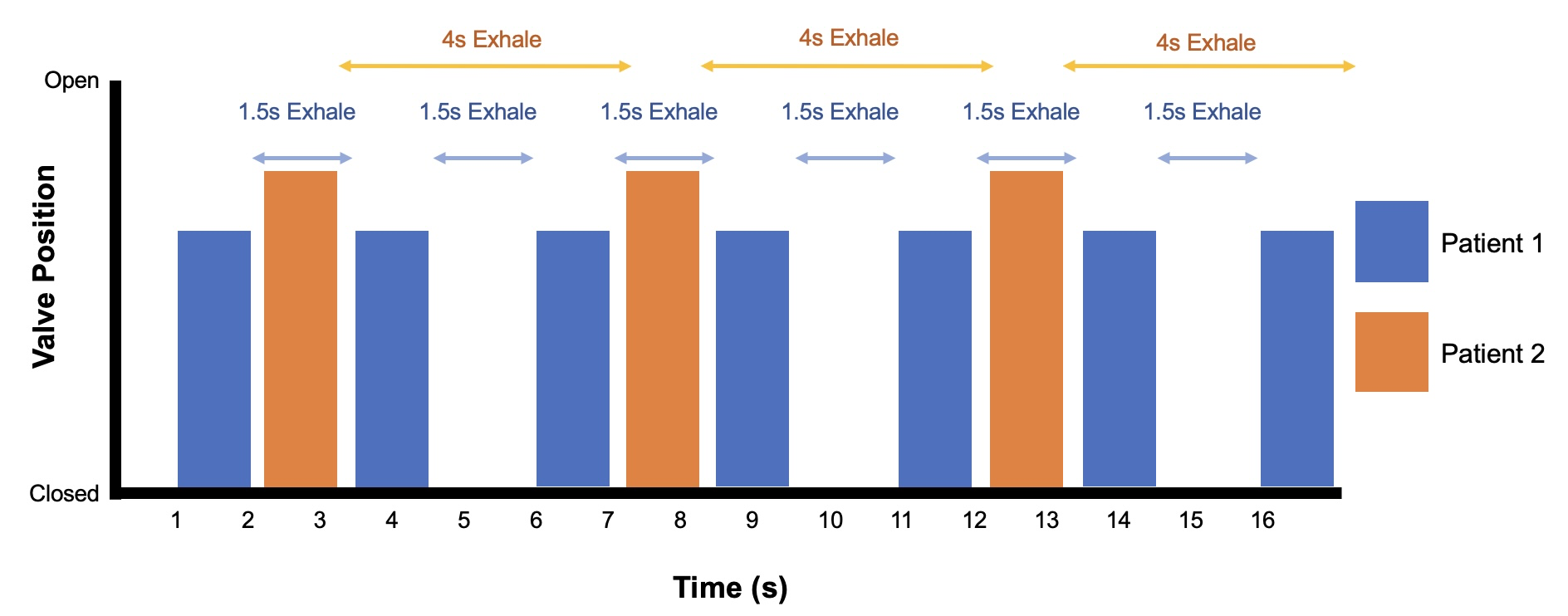

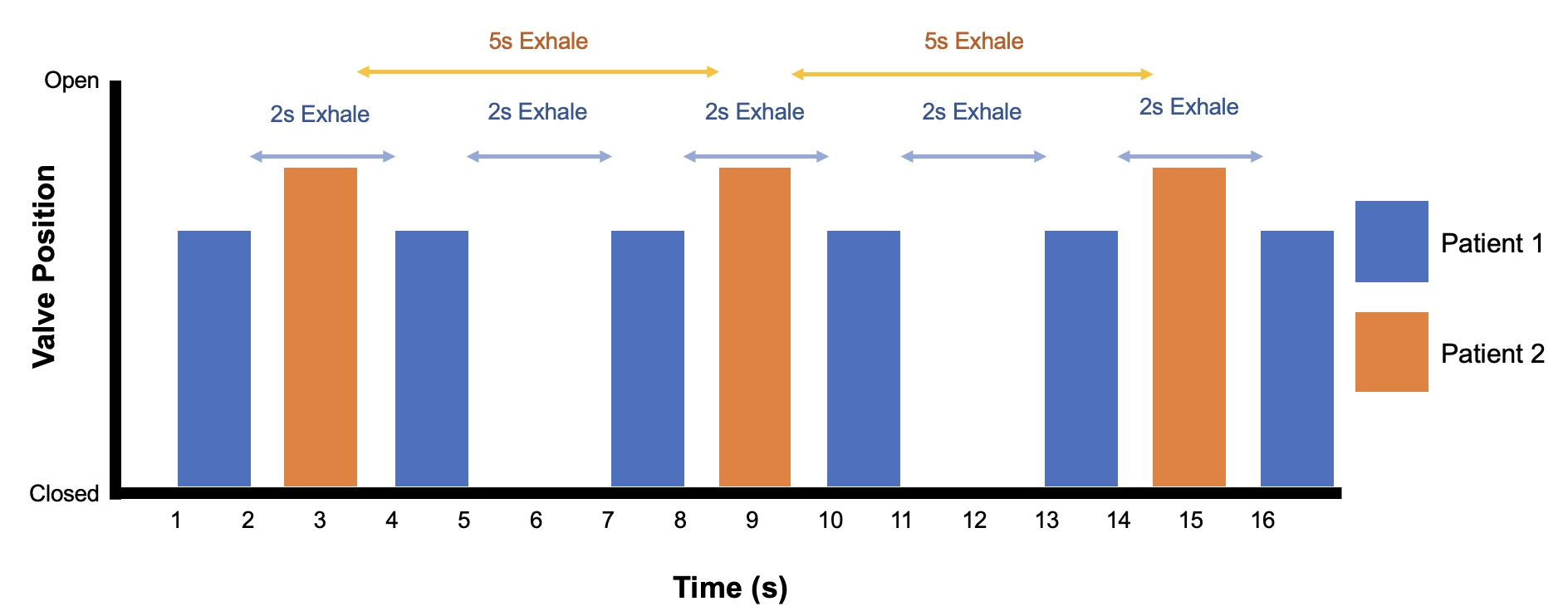
